# Carbapenem Stewardship Program in a Cardiovascular Hospital in Tehran, Iran and Literature Review

**DOI:** 10.1101/2024.12.05.24318537

**Authors:** Elham Nazari, Azin Roumi, Monireh Kamali, Toomaj Sabooteh

## Abstract

**Introduction:** Healthcare-associated infections (HAIs) and antimicrobial resistance (AMR) are critical global challenges. Antimicrobial Stewardship Programs (ASPs) aim to optimize antibiotic use and reduce resistance. This study evaluates carbapenem stewardship compliance in open-heart surgery patients at Shaheed Rajaie Cardiovascular Medical and Research Center, Tehran, emphasizing improved infection control and antibiotic management.

**Methodology:** This descriptive, cross-sectional study assessed carbapenem antibiotic use compliance with national guidelines in open-heart surgery patients at Shaheed Rajaie Cardiovascular Medical Center, Tehran. Seventy patients were selected, and data on demographics, clinical factors, microbiology, and antibiotic use were analyzed using SPSS 26 to evaluate compliance rates and prescribing factors.

**Results:** This study analyzed data from 70 patients undergoing open-heart surgery at a cardiovascular hospital in Tehran. The cohort had an equal gender distribution and a mean age of 59.15 years (SD: 12.0). Of the procedures, 51.43% were elective, while 48.57% were emergencies. The most common surgeries included CABG (45.71%) and valvular procedures (30%). Meropenem was the most prescribed antibiotic (92.9%), with prophylactic use noted in 78.57% intraoperatively. Positive cultures identified Candida albicans (32.7%), Klebsiella pneumoniae (29.8%), and Staphylococcus aureus (22.6%) as predominant pathogens. Serum creatinine levels exceeded the normal range in 55.7% of patients, necessitating dosage adjustments. Antibiotic side effects led to treatment discontinuation in 15.7% of cases.

**Conclusion:** This study highlights a high level of rational carbapenem use in a specialized cardiac surgery ICU, with prescribing guided by infectious disease consultations and adherence to guidelines. Implementing antibiotic stewardship programs can further optimize carbapenem use, reduce antibiotic resistance, and improve treatment outcomes, emphasizing the importance of individualized infection management and monitoring in cardiac surgery patients.

## 1. Introduction

Healthcare-associated infections (HAIs) are a significant challenge for healthcare systems worldwide, as they are associated with prolonged hospital stays, increased treatment costs, elevated mortality rates, and reduced quality of life (1,2). One of the major contributors to HAIs is the inappropriate use of antibiotics. Practices such as incorrect prescriptions, inadequate dosing, or overuse of antibiotics have led to the global crisis of antimicrobial resistance (AMR) (3,4).

The World Health Organization (WHO), in 2011, declared the “Antibiotic Resistance Year” to highlight this issue, warning that AMR drastically reduces the efficacy of treating many infections and could pose an even greater threat to public health in the future. It is estimated that by 2050, approximately 10 million deaths annually could be attributed to AMR (5,6).

In response to this crisis, Antimicrobial Stewardship Programs (ASPs) have been implemented globally. These programs aim to reduce unnecessary antibiotic use, prevent bacterial resistance, and improve clinical outcomes. ASP initiatives typically include monitoring antibiotic prescriptions according to scientific guidelines, consulting infectious disease specialists, and analyzing antibiogram data (7,8).

In Iran, the ASP was officially introduced and implemented in 2017. The program emphasizes restricting the use of antibiotics with a high potential for resistance. Among the antibiotics subject to strict regulation are carbapenems (imipenem and meropenem), colistin, vancomycin, and ciprofloxacin (9).

Patients undergoing open-heart surgery are a high-risk group for HAIs due to their immunocompromised state, the use of invasive devices such as ventilators and catheters, and prolonged hospital stays. These patients are particularly susceptible to severe infections, such as pneumonia and bloodstream infections (10,11). Optimal antibiotic prescription and management are therefore critically important for this population.

At the Shaheed Rajaie Cardiovascular Medical and Research Center in Tehran, the ASP is implemented to control antibiotic use and reduce bacterial resistance. This program enforces restrictions on the prescription of antibiotics such as carbapenems and requires infectious disease specialists’ approval for continuing treatment. ASP forms used in this hospital record patient demographic data, culture and antibiogram results, and drug utilization guidelines (12,13).

This study aimed to assess the compliance of carbapenem (imipenem and meropenem) use with ASP guidelines in patients undergoing open-heart surgery at the Shaheed Rajaie Cardiovascular Medical and Research Center in Tehran. The findings of this research can play a crucial role in improving treatment quality, reducing antimicrobial resistance, and enhancing the optimal management of antimicrobial agents..

## 2. Methodology

### 2.1. Participants and Study Design

This study was a descriptive, cross-sectional research project conducted to assess the compliance of carbapenem antibiotic use (imipenem and meropenem) with the Ministry of Health’s antimicrobial stewardship guidelines in open-heart surgery patients. The methodology was designed and reported in accordance with the STROBE (Strengthening the Reporting of Observational Studies in Epidemiology) guidelines for cross-sectional observational studies to ensure clarity, transparency, and reproducibility. A total of 70 patients (35 males and 35 females) were selected from those admitted to Shaheed Rajaie Cardiovascular Medical and Research Center in Tehran, Iran. These patients had undergone open-heart surgery and received carbapenem antibiotics during their treatment course. The study design followed a retrospective, non-interventional approach, ensuring no influence on patient care while focusing on evaluating prescribing practices and outcomes.

### 2.2. Setting and Variables

The study was conducted in all intensive care units (ICUs) of Shaheed Rajaie Cardiovascular Medical and Research Center, where open-heart surgery patients are treated. Variables assessed in this study included: 1) Demographic information: Age, gender. 2) Clinical parameters: Length of ICU stay, vital signs during prescription. 3) Antibiotic-related data: Type, duration, and dosage of antibiotic use. 4) Medical history: Comorbidities, recent surgeries, and history of antibiotic resistance. 5) Laboratory data: Results of blood, urine, and sputum cultures. The primary outcome of interest was the degree of compliance with the national antimicrobial stewardship guidelines.

### 2.3. Sampling Method

A total of 70 eligible patients were selected using a consecutive, convenience sampling method. The eligible patients were identified from hospital databases within a six-month study period, ensuring adequate representation for analysis.

### 2.4. Inclusion and Exclusion Criteria

Inclusion criteria were Patients aged 18 years or older; Patients who received carbapenem antibiotics (imipenem or meropenem) for at least 72 hours; Patients who survived for at least 96 hours post-surgery. Exclusion criteria were Patients with a known allergy to penicillin or those who developed hypersensitivity reactions to carbapenems; Patients whose antibiotic treatment lasted less than 48 hours; Cases with incomplete medical records or missing clinical data.

### 2.5. Bias

To minimize bias, the study employed multiple measures:

#### Selection bias

Comprehensive screening of all ICU admissions during the study period.

#### Observation bias

Use of a standardized, validated data collection form to extract consistent information.

#### Data quality

Cross-checking collected data with medical records, bedside reviews, and consultations with treating physicians.

### 2.6. Study Size

The sample size was determined based on the total number of eligible patients within the six-month study period. Since no prior nationwide data were available, this study was designed as a pilot project.

### 2.7. Research Method for Data Collection

Data were gathered in three phases, adhering to the STROBE framework:

1. **Identification of Patients:** Using hospital pharmacy records and ICU admission logs to identify those prescribed carbapenems.
2. **Data Extraction:** Reviewing patient medical records to extract relevant demographic, clinical, and microbiological information.
3. **Data Validation:** Cross-verifying extracted data with bedside reviews and staff consultations to ensure accuracy.

The data collection form included sections on patient demographics, antibiotic usage, clinical indicators, and compliance with the Ministry of Health guidelines.

### 2.8. Detailed Explanation of the Research Procedure

Upon ethical approval and obtaining official permission, the research team began by identifying eligible patients through a review of ICU admission logs and pharmacy prescription records. Patients prescribed imipenem or meropenem within the study period were flagged, and their medical files were retrieved for further review. Patients were excluded if: They had incomplete medical records; They were no longer in the hospital or could not be located during follow-up; Data inconsistencies, such as discrepancies in the recorded versus actual antibiotic administration, were observed. For the remaining eligible patients, the research team conducted bedside reviews to collect real-time data, corroborating findings with the nursing staff and treating physicians. This process involved: Documenting demographic and clinical details; Verifying the indications for antibiotic use and its compliance with national guidelines; Recording the results of microbiological tests and any changes in clinical status during antibiotic therapy. Data on antibiotic regimens, including dosages, durations, and indications for prescription, were specifically evaluated against the Ministry of Health’s guidelines for antimicrobial stewardship. Finally, all collected data were anonymized and subjected to statistical analysis to identify trends, compliance rates, and areas for improvement in antibiotic prescribing practices. The results provided actionable insights into optimizing the use of carbapenem antibiotics in the ICU setting.

### 2.9. Validity and Reliability

The data collection tool’s validity was confirmed through expert review by infectious disease specialists and clinical pharmacists. The study adhered to the STROBE guidelines to ensure methodological rigor and reporting quality. All data were reviewed independently by multiple researchers to enhance reliability.

### 2.10. Statistical Analysis

Data were entered into Microsoft Excel and analyzed using SPSS software (version 26). Descriptive statistics (frequencies, percentages, medians, interquartile ranges) were used to summarize qualitative and quantitative variables. Key analyses focused on the degree of compliance with antimicrobial stewardship guidelines and factors influencing deviations.

## 3. Results

In accordance with the national initiative to reduce antibiotic consumption in hospitals and ensure their appropriate use, this study investigated data from 70 patients who underwent open-heart surgery and were hospitalized in the intensive care units of the Shaheed Rajaie Cardiovascular Medical and Research Center. These patients were administered one of the broad-spectrum antibiotics, meropenem or imipenem. All patients who underwent open-heart surgery received general anesthesia and were intubated with mechanical ventilation for at least 8 hours postoperatively. The mean age of the patients was 59.15 years with a standard deviation of 12.0 (ranging from 23 to 92 years).

Table 1 presents the frequency distribution and percentage of demographic and clinical variables of the study patients. As shown, the study population consisted of 35 women (50%) and 35 men (50%). Of these patients, 34 (48.57%) underwent surgery on an emergency basis, while 36 (51.43%) underwent elective surgery. The types of surgery included 32 patients (45.71%) who underwent coronary artery bypass grafting (CABG), 21 patients (30%) who had valvular surgery, and 17 patients (24.29%) who underwent both procedures simultaneously. Additionally, prophylactic antibiotics were administered preoperatively to 4 patients (5.71%), intraoperatively to 55 patients (78.57%), and postoperatively to 11 patients (15.71%) (Table 1).

**Table 1.** Distribution of Frequency and Percentage of Demographic and Clinical Variables of the Studied Patients.

| Variable | N (%) |
| --- | --- |
| Gender |  |
| Female | 35 (50.0%) |
| Male | 35 (50.0%) |
| Surgical Condition |  |
| Emergency | 34 (47.1%) |
| Elective | 36 (51.4%) |
| Type of Surgery |  |
| CABG | 32 (45.7%) |
| Valve-related Issues | 21 (30.0%) |
| Both Surgeries Combined | 17 (24.3%) |
| Antibiotic Prophylaxis |  |
| Preoperative | 4 (5.7%) |
| Intraoperative | 55 (78.6%) |
| Postoperative | 11 (15.7%) |

Table 2 provides information on the distribution of frequency and percentage of underlying diseases, treatment histories, and devices used for the studied patients. As shown, out of the total of 70 patients, 47 (67.1%) had diabetes, 14 (20%) had end-stage renal disease (ESRD), 3 (4.3%) had chronic obstructive pulmonary disease (COPD), and 54 (77.1%) had hypertension (HTN). Additionally, one patient (1.4%) had a history of autoimmune disease, 2 patients (2.9%) had a history of antibiotic use, 8 patients (11.4%) had a history of hospitalization within the past 30 days, and 8 patients (11.4%) had a history of dialysis within the past 30 days. Among the 35 female patients, 2 (5.7%) were pregnant, and 1 (2.9%) was breastfeeding. Furthermore, all patients (70; 100%) were placed on mechanical ventilation with a ventilator, 67 patients (95.7%) had a urinary catheter, 21 patients (30%) used a Sheldon catheter, 68 patients (97.1%) had a central venous catheter (CV line), 69 patients (98.6%) had an arterial catheter (arterial line), and 60 patients (85.7%) had a nasogastric tube (NG tube) (Table 2).

**Table 2.** Distribution of Frequency and Percentage of Underlying Diseases, Medical History, and Devices.

| Variable | Yes<br>N (%) | No<br>N (%) |
| --- | --- | --- |
| Underlying Diseases and Medical History |  |  |
| Diabetes | 47 (67.1%) | 23 (32.9%) |
| ESRD (End-Stage Renal Disease) | 14 (20.0%) | 56 (80.0%) |
| COPD (Chronic Obstructive Pulmonary Disease) | 3 (4.3%) | 67 (95.7%) |
| Hypertension (HTN) | 54 (77.1%) | 16 (22.9%) |
| History of Autoimmune Disease | 1 (1.4%) | 69 (98.6%) |
| History of Antibiotic Use | 2 (2.9%) | 68 (97.1%) |
| History of Hospitalization in the Last 30 Days | 8 (11.4%) | 62 (88.6%) |
| History of Dialysis in the Last 30 Days | 8 (11.4%) | 62 (88.6%) |
| Pregnancy | 2 (5.7%) | 33 (94.3%) |
| Breastfeeding | 1 (2.9%) | 34 (97.1%) |
| Devices |  |  |
| Mechanical Ventilation (Ventilator) | 70 (100%) | 0 (0%) |
| Urinary Catheter | 67 (95.7%) | 3 (4.3%) |
| Sheldon (Sheldon) | 21 (30.0%) | 49 (70.0%) |
| Central Venous Catheter (CV Line) | 68 (97.1%) | 2 (2.9%) |
| Arterial Catheter (Arterial Line) | 69 (98.6%) | 1 (1.4%) |
| Nasogastric Tube (NG Tube) | 60 (85.7%) | 10 (14.3%) |

Table 3 presents the distribution of the frequency and percentage of different antibiotics administered to the patients included in the study. As shown, out of the total 70 participants in this study, 11 patients (15.7%) received colistin, 31 patients (44.3%) received quinolones, 8 patients (11.4%) received aminoglycosides, 11 patients (15.7%) received tetracyclines, 4 patients (5.7%) received imipenem, and 65 patients (92.9%) were prescribed meropenem. On average, each patient received 1.8 antibiotics. The treatment duration for all patients exceeded 72 hours, and all patients received infectious disease consultations (Table 3).

**Table 3.**
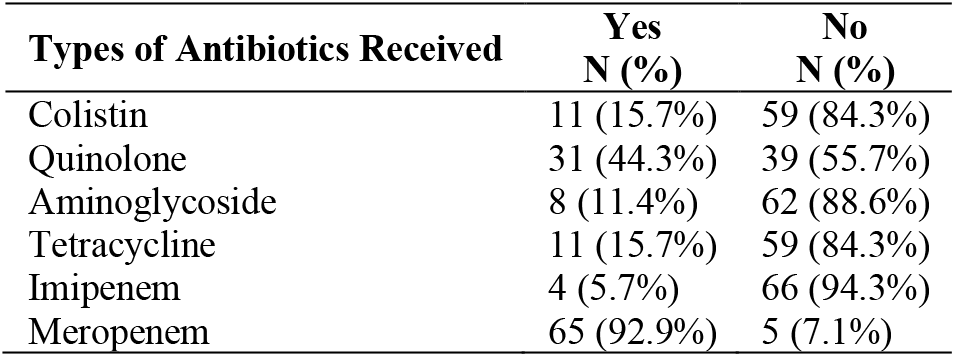
Frequency Distribution and Percentage of Different Types of Antibiotics Received by the Study Patients.

The results of the investigation into the types of cultures performed for patients during their hospitalization revealed that antibiotics were irrationally prescribed for 18.6% of individuals. The most common strains observed in positive cultures were *Candida albicans* (32.7%), *Klebsiella pneumoniae* (29.8%), and *Staphylococcus aureus* (22.6%), in that order. Therefore, the risk of antibiotic resistance to these microorganisms is more significant than for others.

The mean (standard deviation) serum creatinine level of the patients under study was 1.6 (1.1) mg/dl. The lowest and highest serum creatinine levels observed were 0.4 mg/dl and 6 mg/dl, respectively. Given the reference range for serum creatinine (0.6-1.4 mg/dl), it is evident that the mean serum creatinine level in the patients studied is above the normal range. Seventy-two hours after carbapenem administration, 39 patients (55.7%) required dose adjustments due to elevated serum creatinine levels. Additionally, in 11 patients (15.7%), antibiotic use was discontinued due to the occurrence of side effects.

Table 4 presents the data on the frequency distribution and final diagnosis of the type of infection (focus of infection) and the probable focus of infection in the studied patients. As shown, the most common reasons for carbapenem administration were, in order, for the treatment of patients diagnosed with pneumonia (39 patients, 55.7%), sepsis (13 patients, 18.5%), and mediastinitis (11 patients, 15.7%). The most frequent probable foci of infection were the respiratory system (37 patients, 52.8%), bone (12 patients, 17.2%), and bacteremia (10 patients, 14.2%) (Table 4).

**Table 4.** Distribution of Frequency and Percentage of Final Diagnosis of Infection Type (Involved Focus) and Potential Infection Focus.

| Variable | N (%) |
| --- | --- |
| Type of Infection (Involved Focus) |  |
| Pneumonia | 39 (55.7%) |
| Sepsis | 13 (18.5%) |
| Mediastinitis | 11 (15.7%) |
| Urosepsis | 5 (7.1%) |
| Surgical Prophylaxis | 5 (7.1%) |
| Endocarditis | 2 (2.8%) |
| Potential Focus of Infection |  |
| Respiratory System | 37 (52.8%) |
| Bone | 12 (17.2%) |
| Bacteremia | 10 (14.2%) |
| Urinary Tract and Kidney | 5 (7.2%) |
| Central Venous Catheter | 3 (4.3%) |
| Heart | 2 (2.8%) |

Table 5 presents the information regarding the distribution of frequency and percentage of positive and negative cultures from the patients under study, categorized by the site of culture collection. As observed, the majority of cultures were obtained from blood samples of 57 patients (81.4%), sputum samples of 51 patients (72.8%), and urine samples of 47 patients (67.1%). Among these, the highest proportion of positive cultures were found in discharge samples from 9 patients (100%), sputum samples from 40 patients (78.4%), and wound samples from 11 patients (73.3%).

**Table 5.**
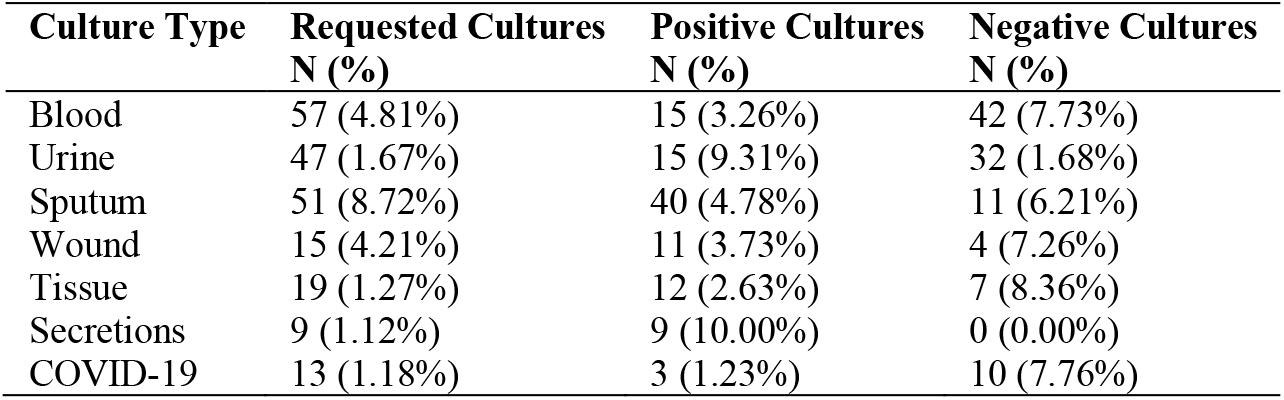
Frequency distribution and percentage of positive and negative cultures in the study patients, categorized by culture site.

Table 6 presents the distribution of frequency and percentage of bacterial species in the positive culture samples from the patients studied, categorized by the sample collection site. As shown, the most prevalent bacterial species in the blood positive culture samples were *Staphylococcus aureus* (5 samples, 33.3%) and *Acinetobacter baumannii* (4 samples, 26.7%). In the urine positive culture samples, *Candida albicans* was most common (11 samples, 73.3%). For the sputum positive culture samples, the predominant species were *Klebsiella pneumoniae* (15 samples, 37.5%) and *Candida albicans* (8 samples, 20%). In the wound positive culture samples, *Staphylococcus aureus* was found in 4 samples (36.4%). In the tissue positive culture samples, *Staphylococcus aureus* was identified in 3 samples (25.0%). Lastly, in the secretion positive culture samples, *Staphylococcus aureus* appeared in 3 samples (33.3%) along with *Escherichia coli* in 3 samples (33.3%).

**Table 6.** Frequency Distribution and Percentage of Bacterial Species in Positive Culture Samples of the Studied Patients, Classified by Culture Site.

| Bacterial Species | Blood<br>N (%) | Urine<br>N (%) | Sputum<br>N (%) | Wound<br>N (%) | Tissue<br>N (%) | Secretions<br>N (%) |
| --- | --- | --- | --- | --- | --- | --- |
| *Staphylococcus aureus* | 5 (33.3%) | 1 (6.7%) |  | 4 (36.4%) | 3 (25.0%) | 3 (33.3%) |
| *Acinetobacter baumannii* | 4 (26.7%) |  | 7 (17.5%) |  | 1 (8.3%) |  |
| *Pseudomonas aeruginosa* | 2 (13.3%) |  | 4 (10.0%) |  | 1 (8.3%) | 1 (1.1%) |
| *Candida albicans* | 2 (13.3%) | 11 (73.3%) | 8 (20.0%) |  | 2 (16.7%) |  |
| *Serratia marcescens* | 1 (6.7%) |  |  |  | 1 (8.3%) |  |
| *Enterococcus faecalis* | 1 (6.7%) |  | 6 (15.0%) | 3 (27.3%) | 1 (8.3%) |  |
| *Klebsiella pneumoniae* |  | 1 (6.7%) | 15 (37.5%) | 1 (9.0%) | 2 (16.7%) | 2 (22.2%) |
| *Escherichia coli* |  | 1 (6.7%) |  | 3 (27.3%) | 1 (8.3%) | 3 (33.3%) |
| *Gram-negative bacilli* |  | 1 (6.7%) |  |  |  |  |
| Total (Positive Cultures) | 15 (26.7%) | 15 (31.9%) | 40 (78.0%) | 11 (73.3%) | 12 (63.3%) | 9 (100.0%) |

## 4. Discussion and Literature Review

The results of the study are discussed in relation to the objectives of the research. The final conclusion is then presented, followed by the application of the findings and recommendations for future research. The current study aimed to evaluate the compliance of carbapenem antibiotic use (imipenem and meropenem) with the standard stewardship guidelines approved by the Ministry of Health and Medical Education of iran in patients undergoing open heart surgery.

### 4.1. Determine the relative frequency of open-heart surgery patients who are empirically administered meropenem and imipenem antibiotics, compared to the standard guidelines approved by the Ministry of Health

In our study, approximately 18.6% of prescriptions were continued without considering culture results or without sending a proper culture for continued treatment, leading to the initiation and continuation of antibiotic therapy. A study conducted by Vazin et al. in September 2017, aimed at investigating the consumption pattern of colistin at Namazi Hospital in Shiraz, showed that only 13% of cases involved empirical therapy. The results of the present study are in many respects consistent with their findings (14).

In a descriptive cross-sectional study by Shoaei and Bagherzadeh, conducted from December 2012 to December 2013, regarding the evaluation of the consumption pattern of imipenem, meropenem, ciprofloxacin, vancomycin, piperacillin, and tazobactam, the findings indicated that 38.5% of prescriptions were correct, while 61.5% were incorrect and empirically prescribed. Most of these incorrect prescriptions were due to the lack of or failure to send patient cultures to the laboratory, leading to the initiation of treatment without considering the relationship between the microorganism and the antibiotic capable of eliminating it (15).

A study by Zakavati et al. in 2019, titled “Review of the prescription pattern of imipenem/cilastatin at Imam Khomeini Hospital in Ardabil,” showed that 64% of patients received imipenem empirically on the first day of hospitalization, and 75.5% of patients were administered imipenem empirically, while only 24.5% of patients had microbiological cultures requested (16).

A retrospective study conducted from September 2014 to February 2015, evaluating the consumption pattern of meropenem at a military hospital, showed that no cultures for antibiotic sensitivity were performed prior to starting meropenem treatment, and 80% of patients received meropenem empirically (17).

In Khoshdel’s study, the antibiotic use patterns of 265 patients were investigated, and the findings indicated that approximately 37% of antibiotic prescriptions were incorrect. The treatment with antibiotics was empirical in 99% of cases, despite the submission of various cultures from the hospital, and sending cultures did not impact the choice of antibiotic (18).

### 4.2. Determining the Cause of Carbapenem Prescriptions in Open-Heart Surgery Patients According to the Approved Guidelines by the Ministry of Health

The most common reason for carbapenem prescriptions in this study was for patients undergoing CABG surgery. The highest rate of antibiotic prescription adherence to treatment guidelines in this study was observed in patients diagnosed with pneumonia (55.7%). In a significant percentage of prescriptions, treatment was initiated based on microbial culture results and antibiotic sensitivity. Additionally, the most common type of pneumonia leading to carbapenem prescriptions was ventilator-associated pneumonia (VAP), which occurs due to prolonged hospitalization and is considered a hospital-acquired infection. VAP is common in post-cardiac surgery patients and has a poor prognosis. It is primarily caused by gram-negative bacteria, predominantly the *Klebsiella pneumoniae* subgroup, and can be influenced by several risk factors such as class IV heart function according to the New York Heart Association (NYHA), pulmonary hypertension, chronic obstructive pulmonary disease, peripheral vascular disease, renal disease, emergency surgery, duration of cardiopulmonary bypass, mechanical ventilation duration, re-intervention, and reintubation. These factors are closely associated with the occurrence of VAP. Patients with VAP showed a significantly higher mortality rate and longer duration of stay in the intensive care unit (ICU) (19).

A study conducted by Vazin and colleagues in September 2017 to investigate the consumption pattern of colistin at Namazi Hospital in Shiraz found that the most common cause of colistin prescription was pneumonia (69%), which is consistent with the findings of our study (14).

Moreover, another study aimed at reducing multi-drug resistance in *Acinetobacter* showed that the limited use of carbapenems could be effective in decreasing the development of multi-drug resistant *Acinetobacter*, which causes ventilator-associated pneumonia (20).

A study conducted in Turkey by Salih and colleagues in 2013 found that the most common reason for antibiotic use was community-acquired infection (57.9%), with pneumonia (20.4%), skin and soft tissue infections (9.11%), and urinary tract infections (7.9%) being the most common infectious diseases (21).

### 4.3. Identification of Microbial Strains Isolated from Open-Heart Surgery Patients Based on the Protocols Approved by the Ministry of Health

The most prevalent strain observed in culture results is *Candida*, followed by *Klebsiella pneumoniae*. This suggests that the risk of resistance to these microorganisms is higher than that of others, posing a greater threat to these patients. Moreover, it is important to note that *Klebsiella pneumoniae* belongs to the Enterobacteriaceae family. Therefore, attention to the resistance of Enterobacteriaceae to the antibiotics imipenem and meropenem will become more critical. Additionally, the rising resistance to *Klebsiella pneumoniae*, which ranks second in the frequency of culture results in this study, has recently drawn attention due to the increasing resistance of *Klebsiella pneumoniae* strains capable of producing carbapenemase. This indicates that these strains have developed resistance to imipenem and meropenem—two widely used broad-spectrum antibiotics in hospitals—and that resistance to these antibiotics is on the rise. In China, the rate of *Escherichia coli* strains producing ESBL was 71% in 2011, and *Klebsiella pneumoniae* ESBL producers exceeded 50%. In India, carbapenem resistance in *Klebsiella pneumoniae* increased from 29% to 57% between 2008 and 2014. These figures show a significant contrast with the 11% resistance rate for *Klebsiella pneumoniae* reported in the United States in 2013 (22).

According to this report, the pattern of antibiotic resistance follows the pattern of antibiotic consumption. Countries with higher antibiotic consumption exhibit a higher prevalence of resistant strains. Antibiotic resistance is a global issue, but antibiotic consumption has the most significant impact on the consuming society (22).

Infections caused by carbapenem-resistant *Klebsiella pneumoniae* are associated with increased mortality in cardiac surgery patients. To reduce mortality rates from these difficult-to-treat resistant bacteria, potential non-reciprocal interventions include maximizing their antibacterial effect and reducing the number of bacterial strains that survive mutations or continue to cause infections (23).

Treating carbapenem-resistant *Klebsiella pneumoniae* infections is challenging, as carbapenems are often considered the last line of treatment for severe *Klebsiella pneumoniae* infections (24).

In 2019, Jaggi and colleagues in India examined the frequency of carbapenem resistance and the prevalence of various carbapenemase genes in *Klebsiella pneumoniae* and *Escherichia coli*. According to the results of this study, carbapenem resistance in *Klebsiella pneumoniae* was significantly higher than in *Escherichia coli* (53.9% versus 15.6%), indicating the need for stricter surveillance in hospital environments (25). In the present study, the most commonly observed strain was *Klebsiella pneumoniae*.

As shown in one study, hospitals with restrictive policies on carbapenem use experienced a reduction in *Pseudomonas aeruginosa* resistance to carbapenems over a 5-year period, leading to a more favorable treatment outcome (26).

According to a study by Wang and colleagues, meropenem resistance was identified as the second most common resistance reported (67.4%). Of 329 *Pseudomonas aeruginosa* samples, the majority were resistant to imipenem (77.5%) and meropenem (64.7%). High resistance of *P. aeruginosa* to imipenem and meropenem is a warning sign for increasing resistance to carbapenems (27).

In their 2015 study, Hong and colleagues suggested that meropenem resistance could vary from 8% to 57%, emphasizing that their prescription and consumption should be carefully managed (28).

David S. Y. Ong and his colleagues conducted a prospective study in 2011 on the relationship between antibiotic consumption and the spread of resistance in *Pseudomonas aeruginosa* and various *Enterobacter* species in the ICU. They concluded that meropenem consumption was associated with the highest risk of resistance in *Pseudomonas aeruginosa*. They also noted that increasing carbapenem consumption leads to increased resistance in beta-lactamase-producing Gram-negative bacteria (29).

In 2007, Ntagiopoulos and colleagues examined the impact of restricting empirical prescriptions of fluoroquinolones and ceftazidime for an 18-month period on the resistance of Gram-negative bacteria in the ICU. Their study showed a significant reduction in resistance to ciprofloxacin in major ICU Gram-negative bacteria after the restriction period (30).

### 4.4. Determining the Average Number of Antibiotics Prescribed in Cardiac Surgery Patients Based on the Approved Guidelines of the Ministry of Health

In the present study, the average number of antibiotics prescribed for patients was 1.87, with one-third of patients receiving at least one antibiotic. Meropenem was prescribed in most patients, followed by quinolone antibiotics, which, in combination with meropenem, were the most commonly prescribed antibiotics. For the majority of patients in this study, prophylactic antibiotics were initiated during the surgical procedure.

A critical assessment of antibiotic use in a hospital in Turkey found that 77% of patients received rational antibiotic therapy, with an average of 1.8 antibiotics per patient, while unnecessary use was reported in 23% of cases. In this observational study, 1350 hospitalized patients were assessed. Of these, 461 (34.1%) received antibiotics for treatment, and 187 (13.9%) received antibiotics for prevention. The indication for antibiotic administration was found in 355 out of 461 treatment patients (77.0%), with an average of 1.8 antibiotics per patient in the treatment group. When antimicrobial therapy was initiated, positive culture results were available for 39 patients (8.5%). Appropriate antibiotic use was observed in 243 patients (52.7%) (21), which aligns with the findings of our study.

In the study by Zhang et al. (2018), antibiotic data were collected from 35 departments in 18 hospitals across 9 provinces. Overall, 67.76% of patients received at least one antibiotic. The most common indication for antibiotics was lower respiratory tract bacterial infections (31).

A study conducted in February 2016 by Gandra et al. (2017) in six hospitals in India reported that 61.5% of hospitalized patients used at least one antibiotic (32).

### 4.5. Determining the Deviation from the Ministry of Health Guidelines for the Prescription of Carbapenem Antibiotics

In the present study, all patients (100%) received infectious disease consultations. Considering that in the majority of patients (81%), carbapenem antibiotic therapy was initiated based on positive cultures, and irrational prescribing was observed in only 18% of patients, the highest level of compliance with the antibiotic prescribing guideline was observed in this study.

A study conducted by Mirzakhani et al., titled *“Review of Antibiotic Prescription in Open Heart Surgery with Stewardship Protocol at the Mazandaran Heart Center”* in 2017, showed that the alignment of antibiotic prescription with the consumption management protocol was at a high level. However, a relatively low adherence to the protocol regarding the number of antibiotics prescribed was observed. This led to reduced long-term efficacy due to microbial resistance and increased costs. In the Shaheed Rajaie Cardiovascular Medical and Research Center, daily consultations and visits by infectious disease specialists take place, ensuring that any antibiotic, even if prescribed by the treating physician, is reviewed and changed by an infectious disease expert. In line with the importance of this issue, a monthly stewardship committee should be formed, which, through the education and cooperation of the entire medical team, can develop an effective strategy to enhance efficiency, reduce microbial resistance, and lower costs. All centers should have infectious disease specialists, with daily infectious disease consultations and visits conducted (13), which aligns closely with the findings of the present study.

A qualitative study on the use of antibiotics in the medical department of a teaching hospital reported irrational antibiotic prescriptions in approximately 23% of cases. The over-prescription of broad-spectrum antibiotics, when a narrow-spectrum antibiotic would suffice, was identified as the most common prescribing error (33).

In a previous study conducted in hospitals in Tehran, the rate of unnecessary or irrational prescriptions was reported to be 0.23%, and the rate of long-term prescriptions was 6.25% (34), which closely approximates the results of the present study.

In a similar study conducted at a university hospital in Chile, 136 patients were studied over a 4-month period. 1.58% of patients received imipenem according to the guideline, while 8.11% of patients were prescribed imipenem without an indication. Imipenem treatment was discontinued in 6.20% of patients by the medical team. Specific measures have been implemented at the Chile hospital for the past four years to reduce the use of imipenem (35).

Carlos A. Diaz Granados, in a 2012 study in Atlanta, showed that a limited antibiotic prescription period in collaboration with infectious disease specialists and clinical pharmacists can play a crucial role in reducing microbial resistance and correcting irrational antibiotic prescriptions. In his study, a total of 992 patients, who were either prescribed broad-spectrum antibiotics or received empiric antibiotic therapy without identifying the pathogen, were included. Targeted antibiotics included imipenem and piperacillin-tazobactam. Treatments were designed based on the guidelines of the Society of Critical Care Medicine, and empiric therapies were restricted. Compared to the control period, there was no significant difference in the mortality rate. However, the length of ICU and hospital stays, as well as the duration of antibiotic therapy, were significantly reduced. Antibiotic resistance significantly decreased, and the correct antibiotic selection significantly increased. Although Diaz Granados did not specify the types of resistant bacteria in his study, which primarily focused on designing a rational antibiotic prescribing model, the results of the present study largely support his findings (36).

In 2011, Y.X. Liew conducted a study in Singapore and reported that 37.8% of carbapenems prescribed in a hospital were irrational, with this issue being more prevalent in the ICU due to concerns about invasive infections. He emphasized the need for antibiotic stewardship programs (ASP) to prevent the growing resistance of carbapenems (37).

An interdisciplinary ASP was implemented in a top hospital in Singapore, where patients receiving broad-spectrum injectable antibiotics were identified daily and subjected to prospective review with immediate feedback. A total of 2084 antibiotic courses were reviewed, of which 24% were inappropriate. Meropenem was most commonly inappropriately prescribed (31.0%). The most frequent reasons for inappropriate use were incorrect selection (51.0%) and incorrect duration (21.4%). Recommendations for discontinuing antibiotics occurred in 34.4% of cases, for dose adjustments in 17.2%, and for changes based on culture results in 86.7% of cases (38).

A study in Croatia reported that irrational antibiotic prescribing was 23%. The most probable reason for improper treatment was the absence of treatment guidelines. Moreover, if a particular antibiotic was overused, the likely cause was the lack of confidence in diagnostic cultures. Improving adherence to published antibiotic treatment guidelines or the distribution of local guidelines will be the most effective tools for reducing this type of misuse. Based on the findings of this study, local antibiotic use guidelines and efforts to promote their implementation will be developed (33).

A study conducted in Turkey by Salih et al. in 2013 found that clinical manifestations of infection, such as the presence of leukocyte counts and prescriptions by infectious disease specialists, were significant positive factors for the appropriate use of antibiotics. As a result, better clinical and laboratory diagnoses, along with consultation with infectious disease specialists, can improve the quality of antibiotic use (21).

### 4.6. Determining the Relative Frequency of Patients Requiring Carbapenem Dose Adjustment Based on the Ministry of Health Guidelines

In the present study, 55.7% of patients (39 individuals) within 72 hours of starting carbapenem treatment required an initial dose adjustment based on their creatinine clearance (creatinine <1.6). Furthermore, 15.7% of the patients had to discontinue the medication due to side effects. Upon reviewing the serum creatinine levels of the patients and considering the normal laboratory range at Shaheed Rajaie Cardiovascular Medical and Research Hospital (0.4-1.6), it was observed that most patients had serum creatinine levels higher than the normal range. Given the cardiovascular involvement in cardiac patients, impaired renal function, as indicated by elevated serum creatinine levels, further complicates the disease process and slows recovery. Additionally, a substantial portion of both imipenem (70%) and meropenem (25%) is excreted unchanged through urine, which necessitates dose adjustment in renal patients.

A previous study conducted in Sudan from September 2014 to February 2015 examined the pattern of meropenem usage in a military hospital. The collected data indicated that 59.2% of patients receiving meropenem required dose adjustments (17), which is very similar to the results of the current study.

In a study by Sakhaiyan et al., the use of imipenem in hospitalized patients undergoing bone marrow transplantation was evaluated, where 35.9% of patients required dose adjustments due to renal failure or low body weight, but dose adjustments were not actually made for them. A total of 64 patients were studied during the research period. Imipenem was empirically started in all patients. In 35 patients (54.7%), the antibiotic was deemed effective. In 51.6% of patients, the duration of antibiotic treatment was found to be inadequate (39).

A study by Shiva in Mazandaran Hospital over three months examined the consumption pattern of imipenem in 100 patients and showed that 64% of the patients had appropriate drug dosing, while 50% adhered to the correct duration of the medication. Only 35.7% of patients, who required dose adjustments, received appropriate dose adjustments. In 83 cases (83%), empirical antibiotic therapy was initiated on the first day of admission, including imipenem in 31 patients (37%). All patients received imipenem as an empirical treatment. The most common antimicrobial drug prescribed alongside imipenem was vancomycin (66 cases). Imipenem was prescribed with consultation from an infectious disease specialist in only 30% of the patients. Fourteen imipenem prescriptions (14%) required dose adjustments based on renal function, but dose adjustments were only made in 5 (35%) cases. Culture testing was conducted for 29 cases (29%). As a result, the high rate of empirical imipenem prescription without considering culture results and antimicrobial susceptibility, the neglect of dose adjustments in renal failure, and the initiation of antimicrobial treatment at the time of admission were identified as the main aspects of irrational imipenem use observed in this study. It is recommended to implement a reliable culture/sensitivity testing and prescribing imipenem based on specific guidelines (40).

## 5. Conclusion

In most patients, the administration of carbapenem antibiotics was initiated based on a positive culture, with irrational prescribing observed only in a few cases. The most common reason for carbapenem administration was for patients undergoing coronary artery bypass graft (CABG) surgery, as well as those diagnosed with pneumonia. The most frequently observed pathogen in the cultures was *Candida*, followed by *Klebsiella*. Additionally, the most prescribed antibiotic was meropenem.

In the present study, all patients had infectious disease consultations, and half of the patients required an adjustment of the initial carbapenem dose based on creatinine clearance. Given that this study was conducted in the cardiac surgery ICU, where patients are typically screened for active infections preoperatively, and considering that due to the specialized nature of Shaheed Rajaie Cardiovascular Medical and Research Hospital, antibiotic prescriptions are made only under the guidance of infectious disease specialists, the results of this study indicate a high percentage of rational carbapenem prescriptions, in accordance with guidelines.

Factors such as aging, diabetes, obesity, and prolonged hospital stays can increase the incidence of hospital-acquired infections in patients who undergo cardiac surgery. Therefore, specialists can partially prevent postoperative infections by accurately identifying and managing factors, including obesity, diabetes, underlying diseases, and the duration of hospitalization, which contribute to infection development.

Additionally, through antibiotic stewardship programs, the overuse and irrational use of antibiotics, such as carbapenems, can be restricted. This approach is an effective solution for preventing the spread of antibiotic resistance and, in turn, plays a significant role in improving infection treatment outcomes and reducing the length of hospital stays.

According to recommendations from reputable medical sources, considering the type of infection and antibiotic sensitivity when prescribing carbapenems significantly increases the likelihood of treatment success under such conditions. Proper management of carbapenem prescribing and use, along with improvements in the quality of this medication’s use, plays a crucial role in improving infection treatment outcomes and reducing mortality rates in the hospital.

### Application of Findings

Forms based on microbiological culture-guided prescribing have been designed, enabling us to use drugs with narrower therapeutic spectra as much as possible. The use of antibiotic stewardship programs is one of the primary global strategies for reducing antibiotic use and controlling the emergence and spread of antibiotic resistance. With an antibiotic stewardship program, prescribing errors for hospitalized patients can be minimized.

Such a program should include training for healthcare staff and regular monitoring of antibiotic prescribing practices. Health policymakers in developed countries have promoted the establishment of mandatory antibiotic prescribing oversight programs in healthcare facilities, and the use of electronic prescribing systems in hospitals, along with feedback on compliance, is considered a key component of any antibiotic prescribing quality improvement program (41).

Appropriate and continuous training based on established guidelines can help enhance physicians’ ability to prescribe suitable antibiotic regimens for patients. Given the continuous changes in various treatment guidelines for infectious diseases, it is essential to develop local treatment protocols for microbial infections in hospitals, based on the latest international scientific evidence, taking into account local microbial resistance patterns and available drugs. Such protocols serve as guidance for specialists and certainly lead to rational antibiotic use, a reduction in microbial resistance, and better therapeutic outcomes. They also increase patient safety by reducing treatment-related side effects, mortality, and healthcare costs.

### Limitations

We cannot definitively discuss the effectiveness of this program based on a short-term study, and it is essential to continue the proper implementation of this program to achieve optimal results. Unfortunately, very few studies have been conducted in this field in our country. However, according to studies from other countries, there is evidence that the implementation of antibiotic stewardship programs has had positive effects on parameters such as reduced mortality, the incidence of recurrent infections, especially respiratory tract infections during the treatment course, as well as the indirect reduction of healthcare costs and the adverse effects of antibiotics. The small sample size and the limitation of this study to a single healthcare center are among the major limitations of this study.

### Suggestions

- Encourage multi-center studies to obtain more accurate statistics and data. Implement antibiotic consumption monitoring programs in the agenda of the drug therapy committees or antibiotic committees of all hospitals nationwide.
- Develop antibiotic prescribing protocols based on the most **common organisms in each hospital and the resistance patterns** of microorganisms, with greater attention given to antibiogram results in the treatment of infections.
- Supervise the rational use of antibiotics and implement established protocols by infectious disease specialists and clinical pharmacists during various stages of antibiotic prescribing and consumption in hospitals.
- In surgical procedures, it is recommended to implement a specific antibiotic consumption monitoring program within the surgical department. For instance, prevention can start with a single dose, and the conditions for increasing the dose or continuing pharmacotherapy post-surgery should be determined based on the type of surgery, surgery duration, and the characteristics of the medication.
- For the proper and effective implementation of antibiotic consumption monitoring programs, cooperation and increased awareness among all hospital healthcare staff are required. This program involves various healthcare professionals such as general physicians, pharmacists, nurses, infectious disease specialists, microbiologists, laboratory staff, and hospital information and technology staff. Inadequate participation from any of these professionals can disrupt monitoring efforts.
- In the long term, during the implementation of the monitoring program, collaboration with experts in the fields of economics and clinical practice is recommended to develop an optimized program, considering both economic and clinical impacts.
- Encourage multi-center studies to obtain more precise statistics and data.
- In addition to cultures from patients, hospital environmental cultures should also be performed. Monotherapy, the primary cause of resistance, should be avoided whenever possible.
- It is suggested that future studies examine the current situation of carbapenem antibiotic consumption compliance (according to the standard antibiotic stewardship protocol) in non-specialized departments and compare this with specialized departments.
- Future studies should also assess the current status of consumption compliance for other commonly used antibiotics in specialized departments, such as colistin (following the standard antibiotic stewardship protocol).

## Data Availability

All data produced in the present study are available upon reasonable request to the authors.

## Conflicts of interest

This article is derived from the master’s thesis in Critical Care Nursing by Ms. Elham Nazari, approved by the Shaheed Rajaie Cardiovascular Medical and Research Center, Tehran, Iran. A portion of the results from this study was previously published as a Persian-language article in the *Cardiovascular Nursing Journal*. Necessary correspondence has been made with the *Cardiovascular Nursing Journal* regarding the publication of this second article, which is written in English and includes a comprehensive literature review, and the required permissions have been obtained. The authors declare that they have no other conflicts of interest related to the content or findings of this manuscript.

## Acknowledgements

We sincerely thank Dr. Behzad Momeni, Dr. Houman Bakhshandeh, Dr. Arash Karimi, Dr. Behzad Yousefi-Yeganeh, Mr. Amin Namdari, and the dedicated staff of all intensive care units at the Shaheed Rajaie Cardiovascular Medical and Research Center in Tehran for their invaluable support and collaboration throughout this study. We are also deeply grateful to the other authorities at this center whose assistance and resources significantly contributed to the completion of this research. Their expertise, commitment, and cooperation have been integral to the success of our work. We are also grateful for the financial support and resources provided by Shaheed Rajaie Cardiovascular Medical and Research Center, without which this research would not have been possible.

## Author Contribution

The preparation of this manuscript involved active contributions from all authors. **E.N**. and **M.K**. oversaw the study design and managed data collection. **A.R**. contributed to data analysis and drafted the initial version of the manuscript. **T.S**. reviewed, revised, and provided comprehensive supervision. All authors reviewed and approved the final manuscript.

## Ethical Considerations

This study was approved by the Medical Ethics Committee of Shaheed Rajaie Cardiovascular Medical and Research Center (approval code: IR.RHC.REC.1400.022). Ethical considerations included: Obtaining informed verbal consent from participants; Ensuring confidentiality and anonymization of participant data throughout the study; Providing participants access to the results upon request; Limiting data usage strictly to the purposes of this research; Adhering to principles of academic integrity and proper citation of all referenced materials. The incorporation of STROBE guidelines ensured the study was ethically and methodologically sound, providing a clear framework for both the research process and reporting.

## Notes

### Competing Interest Statement

The authors have declared no competing interest.

### Author Declarations

This study was approved by the Medical Ethics Committee of Shaheed Rajaie Cardiovascular Medical and Research Center (approval code: IR.RHC.REC.1400.022).

